# CoroNet: A Deep Network Architecture for Semi-Supervised Task-Based Identification of COVID-19 from Chest X-ray Images

**DOI:** 10.1101/2020.04.14.20065722

**Authors:** Shahin Khobahi, Chirag Agarwal, Mojtaba Soltanalian

**Author notes:** Contributed equally to this work (Shahin Khobahi, Chirag Agarwal).

## Abstract

In late 2019, a new Coronavirus disease, referred to as Corona virus disease 2019 (COVID-19), emerged in Wuhan city, Hubei, China, and resulted in a global pandemic—claiming a large number of lives and affecting billions all around the world. The current global standard used in diagnosis of COVID-19 in suspected cases is the real-time polymerase chain reaction (RT-PCR) test. Although the RT-PCR remains the standard reference for diagnosis purposes, it is a time-consuming and expensive test, and moreover, it usually suffers from high rates of false-negatives. Several early works have reported that the sensitivity of the chest Computed Tomography (CT) and the chest X-ray imaging are noticeably greater than that of the RT-PCR test at the initial representations of the disease, making them great candidates for developing new and sophisticated methodologies for analysis and classification of COVID-19 cases. In this paper, we establish the use of a rapid, non-invasive and cost-effective X-ray-based method as a key diagnosis and screening tool for COVID-19 at early and intermediate stages of the disease. To this end, we develop a novel and sophisticated deep learning-based signal and image processing technique as well as classification methodology for analyzing X-ray images specific to COVID-19 disease. Specifically, we consider a *semi-supervised* learning methodology based on AutoEncoders to first extract the infected legions in chest X-ray manifestation of COVID-19 and other Pneumonia-like diseases (as well as healthy cases). Then, we utilize this highly-tailored deep architecture to extract the relevant features specific to each class (i.e., healthy, non-COVID pneumonia, and COVID-19) and train a powerful yet efficient classifier to perform the task of automatic diagnosis. Furthermore, the semi-supervised nature of the proposed framework enables us to efficiently exploit the limited available dataset on COVID-19 while exploiting the vast amount of available X-ray dataset for healthy and non-COVID classes. Moreover, such a semi-supervised approach does not require an expert-annotated lesion area for each class. **Our numerical investigations demonstrate that the proposed framework outperforms the state-of-the-art methods for COVID-19 identification while employing approximately ten times fewer training parameters as compared to other existing methodologies** for classification of the COVID-19 from X-ray images (facilitating efficient training in a limited data regime). We further develop explainable artificial intelligence tools that can explain the diagnosis by using attribution maps while providing an indispensable tool for the radiologist in triage state. We have made the codes of our proposed framework publicly available to the research and healthcare community^1^.

## 1 Introduction

In late 2019, a new disease referred to as Corona virus disease 2019 (COVID-19), emerged in Wuhan city, Hubei, China, and resulted in a global pandemic—claiming a large number of lives and affecting billions all around the world to this date.

Currently, a suspected case needs to be confirmed through the real-time polymerase chain reaction (RT-PCR) test. Although the RT-PCR remains the standard reference for diagnosis, it is a time-consuming and expensive test, and moreover, it usually suffers from high rates of false-negatives. Most notably, several early works have reported that the sensitivity of the chest Computed Tomography (CT) and chest X-ray images is noticeably greater than that of the RT-PCR test at the initial representations of the disease. To fully establish the use of chest CT and X-Ray images as a key diagnosis and screening tool for COVID-19 at early and intermediate stages of the disease, the need for developing novel and sophisticated signal and image processing techniques as well as classification methodologies for analyzing CT images specific to COVID-19 is immediate.

<u>The goal of this paper is to address such a need given that:</u>

- **Early and accurate identification is paramount in controlling the pandemic**. Many research groups are racing to create vaccines and other cures for the new coronavirus. While these efforts look promising, the medical experts warn it make take more than a year before vaccines are ready for practical use. In the absence of such vaccines and cures for COVID-19, it is essential to detect the disease at an early stage and immediately isolate an infected patient from the healthy population. This will support the efforts to *flatten the epidemic curve*.
- **Test kits may not prove as useful as advertised**. With the daily increase in the number of newly diagnosed and suspected cases, the diagnosis has become a growing problem in hospitals around the world due to the insufficient supply of test kits and limited diagnosis rates [1]. With limitations of sample collection and transportation, as well as kit performance, the total positive rate of RT-PCR for throat swab samples has been reported to be about 30 to 60 percent at initial presentation. In the current public health emergency, the low sensitivity of RT-PCR implies that a large number of COVID-19 patients would not be identified quickly and may not receive appropriate treatment or isolation. In addition, given the highly contagious nature of the virus, they carry a risk of infecting a larger population. Moreover, the use of nasal or throat swabs to get at an official diagnosis could be frustrating, particularly when specimens from the upper-respiratory tract might show a negative result even though all clinical signs indicate otherwise. There is therefore an urgent need to look for fast alternative methods that can be used by front-line health care personnel for quickly and accurately diagnosing the disease.
- **CT and X-Ray images are critical in practical diagnosis of COVID-19**. Chest CT and X-ray imaging are among the techniques that are commonly used for the diagnosis of pneumonia-like symptoms of COVID-19, it is relatively straightforward to carry out, and results can be analyzed quickly. A recent study of over 1000 patients discovered that lung computational images were the best method for diagnosing COVID-19 at an early stage and concluded that it should be the primary screening method [2].

COVID-19 causes severe respiratory symptoms and is associated with relatively high ICU admission and mortality. The manifestations of COVID-19 in CT images has unique characteristics, different from the manifestations of other viral pneumonia such as Influenza-A pneumonia [3]. Therefore, there is an untapped potential for the use of CT images in COVID-19 diagnosis.

- **Enhancing the CT and X-ray based diagnosis with artificial intelligence (AI) tools has shown promise**. Some deep learning-based solutions are already developed in China for frontline use to help clinicians detect and monitor the disease more effectively. The outbreak has put significant pressure on imaging departments, that are reading thousands of cases a day. Patients and clinicians typically have to wait a few hours to get the imaging results, but such deep learning tools are improving the diagnosis speed for each case and are helping sites with limited medical resources to immediately screen out suspected patients for further diagnosis and treatment. Given the fact that test kit results are not instant, we must emphasize putting in place a computational imaging-based (e.g., CT or X-ray) procedure that allows more rapid diagnosis whilst limiting the spread of COVID-19. Advanced AI-aided chest CT/X-ray diagnosis systems are urgently needed for accurately confirming suspected cases, screening, and conducting virus surveillance [1].
- **No high-quality public database of CT/X-ray images for COVID-19 patients is currently available**. Considering the fast growth of the disease, public high-quality and well-annotated datasets are non-existent at this point. In fact, even in private or hospital-owned cases where datasets are compiled, the datasets are very limited and still under development. This is exacerbated by limited number of studies and expertise in properly labeling and annotating the existing data, which in turn directly affects the performance of the underlying model both in training and testing phase, making it difficult to train a high-capacity network and to properly assess its performance in real-world applications. As a result, model-based or domain-knowledge-aware methods must be considered to cope with such dataset scarcities as well as deficiencies.
- **Domain knowledge would help with accurate identification when data is scarce**. Problem-specific models play a central role in understanding and design of complex information systems that are common in our age. On the contrary, off-the-shelf data-driven approaches do not need explicit mathematical models and have a wider applicability at the cost of interpretability. Although the data-driven approaches can handle large and complex datasets, they are ignorant to the underlying problem-level reasoning that may be available. Therefore, it is vital to develop a *hybrid* data-driven and domain-knowledge-aware framework to enhance the accuracy and efficiency of deep learning-based COVID-19 diagnosis using CT/X-ray images, while reducing the computational cost as much as possible (we refer an interested reader to consult [4, 5, 6, 7] and the references therein for a detailed explanation of the existing model-based deep learning models).

Note that most existing approaches for classifying COVID-19 cases primarily depend on using pre-trained deep classification networks like ResNet-50 or Inception-v3. One of the main issues with these approaches is that they do not consider the limited dataset of the COVID-19 cases. In addition, these off-the-shelf models are prone to over-fitting issues in a limited dataset regime which is the case for the task of properly detecting the COVID-19 from existing (limited) lung CT/X-ray images. Such an issue arises in deep learning-based models when the capacity of network (number of trainable parameters) is much larger than the amount of information at hand. Moreover, it should be taken into account that there are small differences between the COVID-19 pneumonia and that of other pneumonia cases.

In this work, *we propose a semi-supervised task-based probabilistic model-aware deep learning architectures* for the purpose of chest X-ray image analysis and distinguishing between the healthy, non-COVID infection type, and COVID-19 to assist the radiologist in triage, analysis and assessment of cases associated with the disease. A key aspect of our proposed framework is *the use of AutoEncoders to learn the latent distributions specific to each class as opposed to using off-the-shelf deep neural networks that may be trained using unrelated datasets. The model-inspired and problem-*specific *nature of the networks is expected to increase the identification performance, specifically when data is scarce*. We further develop *explainable AI* tools that can explain the diagnosis by using attribution maps. The proposed methodology can be seen as an amalgamation of supervised and unsupervised learning methodologies in conjunction with transfer learning-based classification, and hence, the resulting architecture enjoys from the benefits of all the aforementioned methodologies and achieves a very high performance in identification of COVID-19 from chest X-Ray images in a multi-label classification task.

## Notation

We use bold lowercase letters for vectors and bold uppercase letters for matrices. (·)^*T*^, (·)^∗^ and (·)^*H*^ denote the vector/matrix transpose, the complex conjugate, and the Hermitian transpose, respectively.

### 1.1 Prior Art

Since the emergence and the global outbreak of the COVID-19, there has been a plethora of research regarding developing automatic deep learning-based diagnosis tools for the classification of the disease using the chest CT and X-ray images. The most notable works so far are as follows. The authors in [3] have established an automatic early screening model for the COVID-19 based on lung CT images. In particular, they considered the problem of distinguishing the COVID-19 pneumonia from Influenza-A viral pneumonia and healthy cases using deep learning techniques. In particular, they used the traditional pre-trained ResNet architecture to first extract the relevant features from the CT images associated with each case, and then, they concatenated an additional fully-connected network with location-attention mechanism to combine both feature extractor and fully-connected classifier network to improve the overall accuracy. The dataset considered in their study consists of total of 528 CT images for training and validation data-sets, with 189 samples of COVID-19, 194 samples for Influenza-A viral pneumonia, and 145 samples for healthy cases. As for the testing dataset, they used a total of 90 CT images where each class represented by 30 samples in the testing dataset. Their studies suggest an overall accuracy of 86.7% for the considered three groups. A similar study on developing deep learning-based automatic segmentation and quantification of COVID-19 was carried out in [8]. Specifically, the authors in [8] proposed a novel deep architecture known as VB-Net to segment COVID-19 related infected regions in the CT images. Furthermore, for preparing the training dataset for their proposed VB-Net, they adopted a human in the loop (HITL) strategy to further enhance the manual annotation of the relevant regions. Finally, their numerical investigations have shown that the resulting VB-Net shows promising results for automatic segmentation of lung CT images specific to COVID-19. This is of particular importance due to the fact their proposed VB-Net can accelerate the development of high quality datasets for the COVID-19 by providing a deep learning-based automatic segmentation technique to accelerate the generation of algorithmic identification of COVID-19. Similar to the work of [3], the authors in [9] utilized a transfer learning methodology to devise a novel deep learning-based algorithm for screening of COVID-19 using lung CT images. More specifically, the authors in [9] first randomly extract regions of interest (ROI) from the CT images. Next, a pre-trained Inception network [10] was utilized in conjunction with a fine-tuning stage for re-purposing the pre-trained Inception network for extracting the features from the corresponding ROIs. Finally, a fully-connected network was utilized to classify the COVID-19 and typical viral pneumonia (a total of two classes). The dataset considered in [10] is consist of 453 CT images of pathogen-confirmed COVID-19 as well as typical viral pneumonia (non-COVID). As for the training dataset, they used 217 CT images for the training and transfer learning purposes. Finally, the proposed model in [10] assumes a total accuracy of 73.1% with specificity of 67% and sensitivity of 74%. Along the same lines, the authors in [11] have considered the identification of COVID-19 based on the chest X-Ray images in lieu of lung CT images. In particular, they considered a transfer learning-based approach using three pre-trained models: ResNet50, InceptionV3, and ResNetV2 for the task of distinguishing between normal and COVID-19 X-Ray images. Their numerical investigation demonstrated that the ResNet50 model achieved the highest accuracy among the considered three pre-trained model with an overall accuracy of 98%. One might notice the very high accuracy obtained in the work of [11]; nevertheless, such a performance is expected due to the fact that the authors have considered a rather simple classification task, i.e. distinguishing between COVID-19 and normal cases. Presumably, a more comprehensive study must consider a rather more significant task such as classification of three classes: healthy, non-COVID and COVID-19 cases or the task of distinguishing between non-COVID pneumonia and COVID-19 cases in that the difference between the manifestation difference of healthy and COVID-19 X-ray images is drastically high and such a task can be performed with high accuracy using even smaller and less complicated models.

The most relevant to our work is the study carried out in [12]. In the mentioned work, the authors consider the development of a deep convolutional neural network architecture for the task of distinguishing between the healthy, non-COVID, and COVID-19 cases using chest X-Ray images. For the training of the resulting network known as COVID-Net, the authors prepared an open publicly available dataset referred to as COVIDx dataset which is an amalgamation of two open access data repositories: the COVID-19 image data collection and the RSNA Pneumonia Detection Challenge dataset [13, 14]. The COVIDx dataset consists of 16, 756 chest radiography samples with 76 radiography images for the COVID-19 case, 8066 images for the healthy patients, and 5526 patient cases who have non-COVID pneumonia. As for the development of the COVID-Net proposed in [12], the authors utilized a human-machine collaborative design strategy to prototype the network in conjunction with network architecture exploration design tool provided by the Darwin AI, Inc. Finally, it was shown that the COVID-Net architecture achieves an overall accuracy of 92.4% with a sensitivity of 95%, 91% and 80% for healthy, non-COVID, and COVID-19 classes, respectively. Moreover, the achieved positive predicted value (PPV) for each class is as follows: 91.3% for the healthy class, 93.8% for the non-COVID infection type, and 88.9% for the COVID-19 case.

### 1.2 From Conventional to Task-Based Deep Learning

A conventional AI, and specifically deep learning, solution for medical applications entails the following stages:

- **Data Preparation**: During this phase, a large amount of data from some predefined categories will be collected. In particular, in the case of detecting the COVID-19 from chest radiography imaging, one need to prepare a dataset consisting of not only COVID-19 cases, but also healthy cases as well as presumably similar viral pneumonia-like cases that resemble similar manifestations of lung CT. Such a comprehensive dataset further enables the model to properly distinguish between the cases resulting in a more accurate classification. Next, the collected data is required to be annotated by an expert for preparing the labels and ground-truth data for the next steps.
- **Model Design and Training**: At this stage, one seeks to design a deep learning-based framework suitable for the underlying task at hand, i.e. classification of the COVID-19 cases from the chest radiography imaging (e.g., X-ray or CT imaging). This step conventionally consists of designing an image processing unit to first extract the useful features of likely high-dimensional input signals (images). Then, a classifier network will be trained to distinguish between different classes according to the learned features. Note that the infection regions in chest radiography images are low-contrast and admit a large variation of shape and position in different suspected cases, and hence, pinpointing the infected region in the images is an extremely challenging task in designing a successful image processing unit. However, with the advent of convolutional neural networks (CNNs) and the more recent 3-D CNN models (suitable for analyzing CT images), one can easily tackle such a difficult task when the input data is likely high dimensional. Indeed, the CNN model has shown a great promise in medical imaging applications and currently is the de facto model for such tasks. In this paper, we also make use of CNN models to design a powerful classifier as well as a novel image analysis methodology. After a successful design of the model, one seeks to learn the network parameters using the prepared dataset at the previous stage. Another challenge in this stage is that each class needs to be well-represented (i.e., a large number of data is required for each class) so the network can generalize well to unseen cases.
- **Testing**: Finally, during this stage the model will be tested on data points that have not been shown to the model during the previous training stage, where one examines the performance of the model in predicting new cases and investigate whether the model generalizes well or not. Note that the testing and training stages are closely intertwined and a successful training might not result in a good performance in testing stage. For instance, if the network has a large number of parameters to be optimized (i.e., very high capacity) and the training data points are limited, the model is susceptible to over-fitting, leading to a small training error and a large test error. In such a case, one needs to either increase the available data at hand via either collecting new samples or using advanced data augmentation techniques, or to change the underlying model entirely.

In light of the above, and in the case of properly classifying the new COVID-19 virus from chest X-ray images, there exist several challenges that one might face in all of the aforementioned stages: In the case of the new COVID-19 virus, there is a lack of high quality and well annotated datasets. In fact, the studies as well as the expertise required in properly labeling and annotating the existing data points barely exist for such efforts. This in turn directly affect the performance of the underlying model both in training and testing phase—making it difficult to train a high capacity network and to properly asses its performance in real-world clinical applications. On the other hand, there exist a vast amount of information and dataset on other pneumonia-related diseases (e.g., Influenza-A) and one needs to design a sophisticated model that can deal with incorporation of other *large* datasets with the existing *limited* data on COVID-19, so that both classes can be well-represented while training the network. Note that such a mixture of limited representation of one class and another well-represented class can easily degrade the performance of a classifier. Hence, it is of paramount importance to develop novel sampling techniques for training purposes. Second, due to the limited amount of training datasets on the new corona virus, one should avoid using high capacity networks in that they are highly prone to over-fitting for limited data. One way to alleviate this problem is to use the transfer learning method. In transfer learning methodology, one takes a pre-trained network that can successfully perform a task in another context, and transfer the model using a fine-tuning stage to the new task. Although such an approach can be undertaken in the case of COVID-19 classification, the performance of the resulting network can be limited due to the fact that the pre-trained network might not be able to extract the unique features entirely, as it is trained on another dataset. Consequently, it is of importance to design a novel hybrid deep-learning solution for this problem so one can be able not only to harness the power of deep neural networks but also be able to take into account the underlying distribution of the data and design a highly tailored model that allows for the extraction of unique features of each class for this specific task, and does it efficiently.

As mentioned before, most of the existing approaches for classifying COVID-19 cases primarily depend on using a single pre-trained deep classification networks like ResNet-50 or Inception-v3 to perform the underlying task. On the other hand, a common narrative among all the existing works is the fact that the algorithm and the network is designed in a supervised manner (i.e., supervised learning methodology), where the underlying model makes use of expert knowledge and heuristics for both extracting the relevant features from the data and the network architecture design. Such a supervised methodology requires a human in the loop strategy for most (if not all) of the stages in the model and further has an inherent bias towards the way it is being supervised which in turn will reduce the possibility of letting the model exploit unique information from the available limited data at hand during the training (that might seem irrelevant to the supervisor). In order to tackle the problems associated with supervised learning methodology, one can resort to unsupervised and semi-supervised learning methods. Specifically, in an unsupervised learning framework, one designs a model that can intelligently and automatically gain insight from the raw data without supervision and further use the derived information for the task of decision-making and classification. In fact, all of the existing works on identification of COVID-19 from medical radiography images make use of pre-trained networks for preparing the data to be propagated through the classification network in a supervised manner. These models are often ignorant to the fact that the classification is not the only purpose of the underlying algorithm and there exist other stages that can dramatically affect the performance of the model. In particular, an unsupervised learning methodology for compression, feature extraction, and denoising at early stages of the network can result in a tailored task-specific understanding of the data which can greatly improve the performance of the classification algorithm. On the other hand, a semi-supervised methodology can be interpreted as a bridge between supervised and unsupervised learning approaches. In a semi-supervised approach the classification task is performed in a supervised form, while learning the representation of the data is performed in an unsupervised manner to allow for an enhanced generalization of the classifier.

In light of the above discussion, we will use a semi-supervised methodology to design a highly tailored deep learning-based framework to harness as much information as possible from the limited available information at hand. Consequently, we propose a model that first captures and learns the distribution and the latent representation of each case by using AutoEncoders (AEs) (in an unsupervised manner). Once the underlying distributions (and correspondingly, the *unique* features of each case) are learned, we use a feature-aware classifier to properly detect the different cases, including the COVID-19 from the chest X-ray images, in a supervised manner. Such a methodology differs from the existing deep learning models for the task of COVID-19 classification as our model ensures learning the unique data and feature distribution of Healthy and non-COVID cases independently. Namely, the current methodologies use a pre-trained feature extractor network that is trained on non-related datasets (e.g., real-world images) and usually have a very large number of parameters for fine-tuning. Moreover, they might not be suitable for extracting intricate features specific to COVID-19 and other pneumonia cases as they are not trained on these datasets.

An important question in a classification task using deep-learning models in healthcare applications is whether one can asses the decision of the model. In this work, we further seek to develop an intuitive model-aware deep learning framework that facilitates interpreting different parts of the network as well as its decision using the *attribution mapping* methodology. This will allow for further analysis of the data and hopefully can be utilized by the healthcare professionals for another level of scrutiny on the diagnosis made by the framework. We refer the interested reader to our previous work [15] and the references therein for a detailed explanation of the attribution mapping optimization technique used in the context of explainable machine learning. Last but not the least, the proposed methodology can be directly used for data-augmentation purposes to increase the generalization performance of the underlying model. In other words, the trained AEs can be used to sample the underlying captured distribution of each class for data augmentation purposes—an approach we will consider in a future publication on this topic.

## 2 Proposed Semi-Supervised Methodology

In this section, we present the design paradigm behind the proposed deep architecture. As mentioned before, we pursue the design of an intuitive hybrid semi-supervised deep learning-based solution for the task of early screening of COVID-19 from chest X-ray images that can address the problems covered in the previous sections. Specifically, we make use of the recently released COVIDx dataset [12] and consider a semi-supervised learning approach in designing a two stage deep learning-based algorithm for distinguishing between the 3 classes of Healthy, non-COVID pneumonia, and COVID-19 infection. In the sequel, we refer to the non-COVID pneumonia class simply as *Pneumonia* or *non-COVID*.

The proposed methodology is comprised of the following two modules: 1) the **T**ask-Base **F**eature **E**xtraction **N**etwork (TFEN), and 2) the **C**ovid-19 **I**dentification **N**etwrok (CIN). As mentioned before, the majority of existing works are focused on extracting the relevant features from the medical radiology images (either CT or X-ray images) using pre-trained networks which are usually trained on real-world images and might not be able to extract the intricate features related to medical images and specific to our problem. Hence, to address this issue, we propose to use a task-specific feature extraction network that is tailored to handle chest X-ray images associated with the three classes considered in this work, i.e. Healthy, non-COVID and COVID-19 cases. Furthermore, due to the closeness of the chest X-ray manifestation of COVID-19 as compared to to other pneumonia diseases, the proposed TFEN module will designed in a manner to signify the abnormality regions in the chest that is most affected by COVID-19 as compared to healthy and non-COVID classes in a semi-supervised manner. Namely, we make use of AutoEncoders in this module and train the overall TFEN module in a semi-supervised manner to extract the relevant features and information from the raw data. The next step in our proposed methodology, is the CIN module which is tasked to perform the classification based on the extracted residual features from the TFEN module. As for the CIN module, we make use of transfer learning strategy to train a powerful classification network. In the following parts, we give a detail description of each module.

### 2.1 TFEN Module Architecture

In this part, we consider the design of a network that allows for segmentation of abnormal regions in the chest X-ray images that appear due to the existence of either non-COVID or COVID-19 pneumonia. Then, such an abnormal regions will be fed into the classifier to differentiate between the three classes. It is a well-known that the success of supervised learning approaches in most classification tasks is closely related to the existence of high-quality datasets with a very large number of data samples. However, this is not the case in the classification of the COVID-19 from the X-ray images, as the available datasets are either not freely available to the research community, or comprised of very limited cases of COVID-19. Hence, any deep architecture with very high number of parameters trained on these datasets are prone to over-fitting and other classification issues. Furthermore, there exist very high-quality datasets on healthy chest X-ray images as well as non-COVID pneumonia cases. Note that human professionals can detect and segment the anomalies appearing in the chest X-ray images by training on very limited data points, and can further generalize the gained prior information to unseen cases. Namely, by providing a human professional with limited healthy and pneumonia chest X-ray images, the person can easily detect the abnormal regions and differentiate between the two very accurately. Indeed, humans make use of *prior information* very efficiently to recognize the abnormal sections of the X-ray images. This hints us to develop a semi-supervised feature extraction network that allows for obtaining the relevant *prior information* necessary for anomaly detection in the chest X-ray images, and then, exploit this *prior* to perform the classification task in order to mimic the human behaviour more closely. Due to the fact that the COVID-19 samples in the considered dataset are very scarce as compared to the other two classes (e.g., Healthy and non-COVID infection), we first use the information from these two high quality classes (Healthy and non-COVID) as a basis for detecting the abnormal regions in the chest X-ray images that will be used as the input to the classifier network.

Accordingly, we equip the network with a deep sub-architecture, known as AutoEncoder (AE) in the literature, that allows it to first capture the relevant latent representation of the Healthy and non-COVID classes. This module is tasked with learning the *prior* information necessary to extract unique features related to segmenting the the abnormal regions. Once this segmentation is extracted, the network will be equipped with a classifier deep architecture to accurately distinguish between the three classes based on the extracted abnormal regions; more on this below.

Broadly speaking, an autoencoder system is a generative model comprised of an encoder and a decoder module that are sequentially connected together. The goal of an AE module is to learn an abstract latent representation (the relevant features/basis-vectors) of the input data while providing a system to reconstruct the data from the learned abstract representations. Consequently, the input to such a system is a set of signals following a certain distribution, i.e. ***x*** ∼ 𝒫(***x***), and the output is the recovered signal 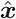 from the decoder module using the latent representations. In short, the goal is to jointly learn an abstract representation of the underlying distribution of the signals through the encoder module, and simultaneously, learning a decoder module allowing for reconstruction of the input signals from the obtained abstract representations. Therefore, an AE module can be defined by two main functions: *i*) an encoder function *ε*_**Φ**_ : *χ* ↦ *Ƶ* parametrized on a set of trainable variables **Φ**, that maps the input signal ***x*** ∈ *χ* into a new vector space known as the feature or latent space *Ƶ*, and *ii*) a decoder function 𝒟_**ψ**_ : *Ƶ* ↦ *χ* parametrized on **ψ**, which maps the data from the latent space *Ƶ* back into the original signal space *χ* Hence, the governing dynamics of a general AE can be expressed as

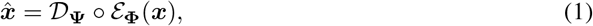

where 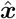 denotes the reconstructed signal. The learning of such an autoencoder system is usually carried out by minimizing the reconstruction error between the input and output, viz.

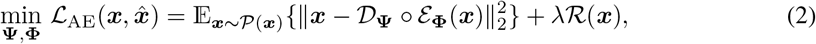

where *λ* > 0 is the regularization coefficient, and ℛ (***x***) denotes a regularization function to assist the learning process that provides domain-knowledge for the training. Indeed, by constraining the latent representation both in dimension and structure (presumably different from the input dimensions), the network can learn and discover unique patterns from the data (i.e., gaining the prior information/basis for reconstructing the input farther in the network). The specific choice of the structure of the latent variables ***z*** = *ε*_**Φ**_ (***x***), and the deep architecture used for the decoder and encoder module gives different flavours of autoencoder systems.

In this work, we consider the learning of task-specific features using two independent autoencoders. Following the discussion on how humans exploit the prior knowledge to efficiently distinguish between different classes by seeing limited data, we consider assigning one autoencoder module to each class of Healthy and non-COVID cases (a total of two autoencoders). Namely, we train two AE systems to extract the relevant features of the two classes of Healthy and non-COVID, then, we use these autoencoders for identification of the abnormalities appearing in infected chest X-ray manifestations and use that information for training the classifier module. The structure of the proposed TFEN module is thus as follows. Let 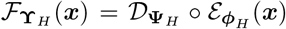 and 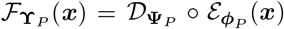 denote the assigned AEs to the Healthy and non-COVID (pneumonia) classes, respectively. Then, we train each AE to learn the relevant features of each class by showing each AE system only the information for the underlying class assigned to them. Let *χ*_H_ and *χ*_P_ denotes the set of sample point in the dataset corresponding to the Healthy and non-COVID chest X-ray images with a cardinality of *N*_*H*_ = |*χ*_*H*_| and *N*_*P*_ = |*χ*_*P*_|, respectively. Then, we consider the problem of learning each AE sub-system according to the following optimization problem:

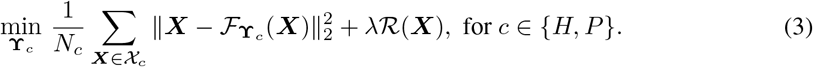

The regularization term ℛ (***X***) in (3) is defined as follows. Due to the fact that the goal of the TFEN module is to first gain some prior knowledge in terms of feature representation of each considered class through the corresponding autoencoders to extract the abnormal regions in the chest, it is desirable to further constrain the learning of latent representation of each class ***c*** ∈{*H, P*} to be distant from of each other in the latent space. Note that our goal is to identify the three classes considered in this work based on the anomalies appear in the chest X-ray. Let 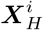 and 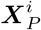 denote two samples from the X-ray images corresponding to Healthy and non-COVID classes, respectively. Then, the anomalies appearing in the infected chest manifestation appear as local variations in certain parts of the healthy chest. Furthermore, let us consider the *ℓ*_2_ norm as a measure of distance between a Healthy and non-COVID chest X-ray image quantifying the affected regions. Then, it might be the case that there exists another healthy X-ray image 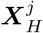 such that, 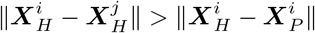. Such a scenario is very likely to happen in our problem, as differentiating between Healthy, non-COVID and COVID-19 might depend on very small variations in the chest X-ray images, which will considerably impact the performance of the classifier. In order to alleviate this problem, i.e., to avoid two different classes mapped into similar latent representations, we utilize the following regularization function acting on the latent variables 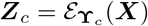 during the training:

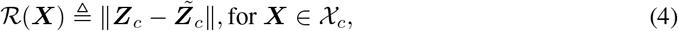

where 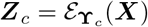 and 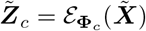 with 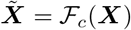 denoting the output of the autoencoder assigned to class *c* ∈ {*H, P*} (i.e., the reconstructed signal). In other words, the above regularization term imposes the constraint that the reconstructed image and the input image must be mapped close to each other in the latent space. Such a regularization term has been successfully used in other deep learning-based algorithms in medical imaging applications and has shown to significantly improve the performance of the underlying model (e.g., see [16, 17] and the references therein).

After the successful training of each individual AE system, we are equipped with a system that is capable of representing the underlying latent representation of Healthy and non-COVID pneumonia cases through the two AE sub-systems. Now, we discuss the next stage in the TFEN module. Let ***X*** denote any data-point from the underlying dataset with all three classes represented. For instance, let us consider the case that ***X*** is a sample from the non-COVID class. Further let 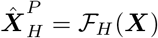 and 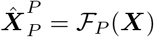 denotes the reconstructed signals when ***X*** (belonging to non-COVID class) is fed to the two AEs, respectively. An interesting observation here is that the AE trained on Healthy cases will not be able to reconstruct the input fully as the class of the input (non-COVID in this example) is different from the class assigned to the AE. On the other hand, when a sample from non-COVID case is fed into the AE corresponding to non-COVID cases, the difference between the reconstructed image is much smaller than that of the Healthy autoencoder. In other words, the Healthy autoencoder tries to create a healthy-looking chest X-ray image while signifying the anomalies appearing due to the non-COVID infection regions. Hence, one can capture those anomaly and differences between different classes by simply considering the absolute intensity differences between the reconstructed signals and the input of each individual AE, i.e. the residual images 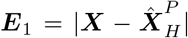 and 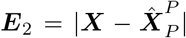. The same argument holds for the case that the input is sampled from COVID-19 data points. Indeed, the residual images ***E***_1_ and ***E***_2_ are providing a measure of distance between the input sample and the corresponding classes that each individual AE is trained on. With this observation in mind, we suggest to use the residual images as the output of the TFEN module to be fed into the classifier network. More concretely, we define the output of the TFEN module as the 3D tensor 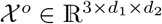 whose horizontal slices are given by:

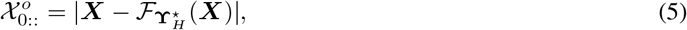

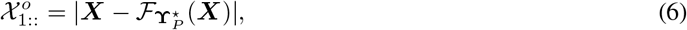

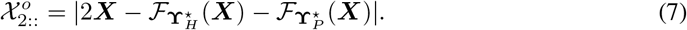

Briefly speaking, we first train the TFEN module comprising of two individual autoencoders assigned to the classes of Healthy and non-COVID cases. The training is performed independently from the classifier network and in a semi-supervised manner to allow for extracting task-specific features related to the classes. Next, after the training of the AEs is completed, we treat the TFEN module as an stand-alone stage providing the classifier with the tensor of residual images. Finally, we train the classifier based on the output of the TFEN module.

One may raise the question that instead of considering two AEs, we could have considered 3 AEs assigned to all three classes. We stress again that, in the case of identifying COVID-19 from X-ray images, the amount of data samples for COVID-19 is extremely scarce and is not sufficient to learn useful latent representations of the chest manifestation of the underlying class. Instead, we inject the information regarding the COVID-19 to the algorithm pipe-line during the training of the classifier. In fact, for training (and also inference), each point in the dataset will be cross-correlated with the two AEs in the TFEN module successfully trained on two classes of Healthy and non-COVID (which we have enough samples to capture the relevant features), and the output is the 3 dimensional tensor consisting of residual images to perform the classification task accordingly.

### 2.2 CIN Module Architecture

In this section, we present our classification network, which we refer to as the CIN module. The underlying task of a classifier model is to map a given (likely high-dimensional) input to one of the corresponding classes, i.e. performing the mapping *χ* ↦ 𝒞, where 𝒞 denotes the space of desirable classes. In this work, we consider the classification of Healthy, non-COVID pneumonia, and COVID-19 from the chest X-ray images, i.e. we consider 𝒞 = {Healthy, non-COVID pneumonia, COVID-19}. As discussed in the previous part, we use the output of the TFEN module as the input to the classifier network. The output of the TFEN module is a 3D tensor that contains the information regarding the residual images obtained from each already trained AEs. Namely, given a sample point from the dataset (representing all classes), we first feed that image into the two trained AEs corresponding to Healthy and non-COVID classes. Then, we construct the residual tensor *χ*° according to (5)-(7). Next, the residual tensor is fed to the classifier for identifying the underlying class of the input image.

For designing the CIN module, we make use of the convolutional neural networks (CNNs) and the idea of transfer learning. Transfer learning is a methodology concerned with transferring knowledge from a network that is trained on another (but similar) task to perform a different task that is similar in nature. In particular, during the transfer learning, one consider the use of a pre-trained deep architecture that is trained on a very large dataset, and fine-tune the weights of the pre-trained network, to transfer the pre-acquired knowledge (in terms of fine-tuning the weights of the network) to perform another task. In particular, a pre-trained CNN on image analysis task is shown to be able to extract the significant patterns appearing in images. Hence, due to the nature of our work that deals with X-ray images, it is most suitable to consider a pre-trained network on images for fine-tuning for our purpose. Furthermore, it is well-known that the use of pre-trained network for transfer-learning not only reduces the training cost, but also immensely helps the performance in the case of limited datasets. Due to the success of transfer learning methodology and the promising fact that the pre-trained networks (if used properly) can provide superior performance for classification tasks, in this work we consider a modified light-weight ResNet-18 architecture pre-trained on ImageNet [18] for our CIN module to perform the task of classification.

As for training of the CIN module, we first feed each data sample in the COVIDx dataset into the TFEN module, and construct the output tensor consisting of three residual images as discussed before, and we perform the training of the our classifier network based on the received output of the TFEN module and the corresponding classes. Furthermore, note that the COVIDx dataset is imbalanced, *i*.*e*. the data points for each class is different than others, and hence, we consider the training of the CIN module using a class weighted entropy loss function (see Sec. 3.2 for more details). Figure 1 demonstrates a visualization of the overall proposed methodology.

**Figure 1:**
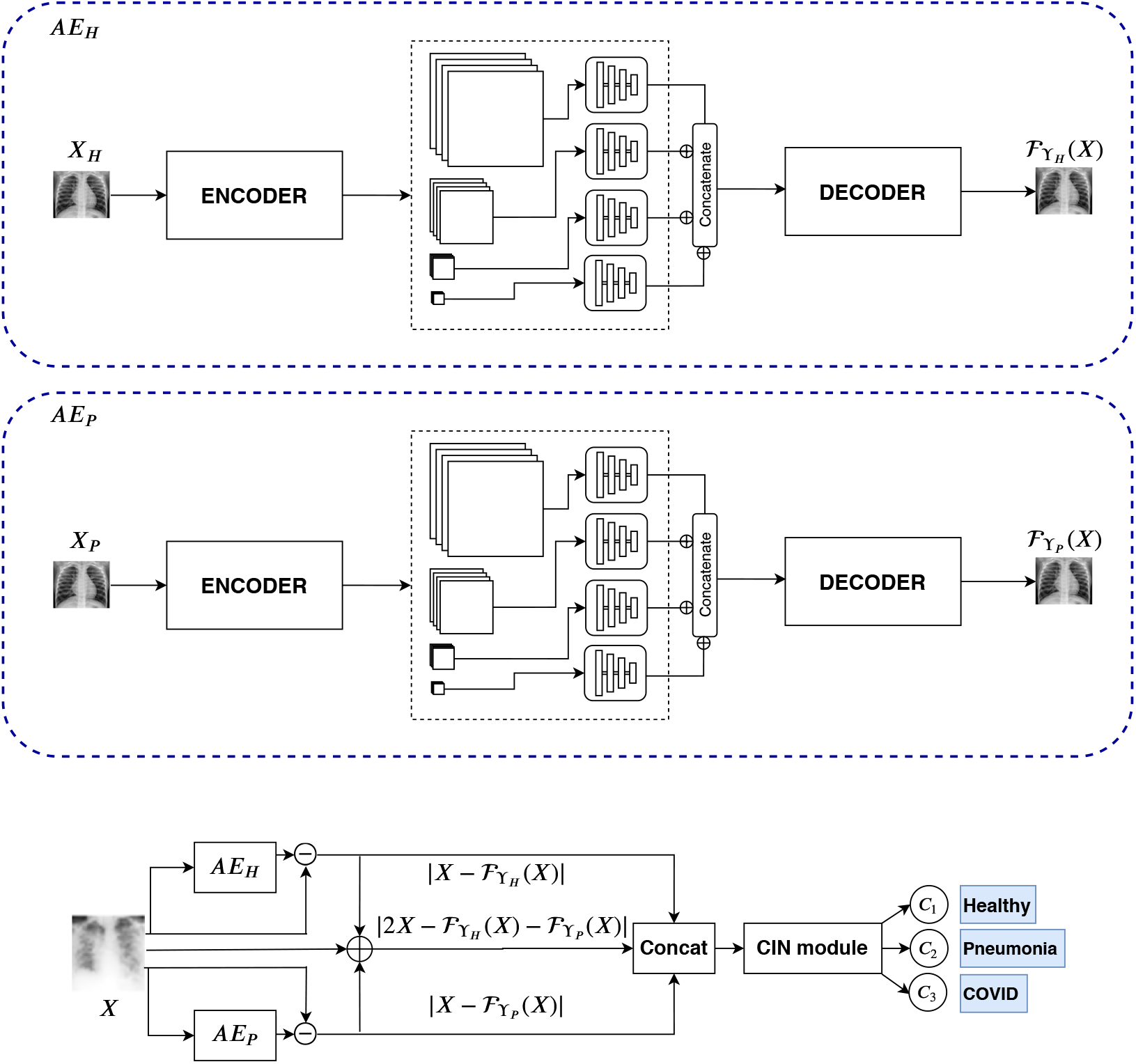
Proposed classification model for Chest X-ray images. AE_*H*_ and AE_*P*_ are two AutoEn-coders trained independently on the images from Healthy (*H*) and non-COVID Pneumonia (*P*) patients respectively. The CIN module, a pre-trained ResNet-18 network (See Sec. 3.2 for details), is trained on the residual images (pixel-wise absolute intensity difference) between input image and the reconstructed image from the AutoEncoders.

## 3 Numerical Results

### 3.1 Dataset

For quantitative and qualitative evaluations, we used the COVIDx dataset [12] which is a combination of two different publicly available datasets, *i*.*e*., some limited COVID-19 image data collections [13], and RSNA Pneumonia detection dataset [14]. We generated the training and testing dataset following the work of [12]. The total number of training and testing images in our dataset were 16576 and 1953 respectively. In Table 1, we provide the number of training and testing images present in the individual categories.

**Table 1:**
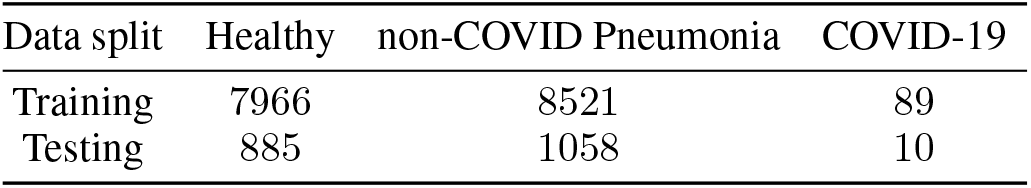
COVIDx dataset [12] image distribution for individual categories.

| Data split | Healthy | non-COVID Pneumonia | COVID-19 |
| --- | --- | --- | --- |
| Training | 7966 | 8521 | 89 |
| Testing | 885 | 1058 | 10 |

### 3.2 Training and Implementation Details

As discussed, our proposed model comprises of two modules, *i*.*e*., the TFEN and CIN module. We use a Feature Pyramid based AutoEncoder (FPAE) [19] network architecture for independently learning the individual distribution of Healthy and non-COVID pneumonia Chest X-ray images in the TFEN module. The feature pyramid framework for designing the autoencoders enables learning a latent representation by concatenating the features from different resolutions. The FPAE consists of a 7-layer convolutional Encoder and Decoder blocks and the classification network is a pre-trained ResNet-18 convolutional network. The FPAE and classification model were trained using Adam optimizer [20] for 50 and 10 epochs using a learning rate of 10^−4^ and 10^−3^, respectively. We use horizontal and vertical flips as data augmentation while only training the CIN module (i.e., the TFEN module is trained without data-augmentation). Our models were implemented using the PyTorch [21] deep learning library. We first performed the training of the proposed TFEN module on the Healthy and non-COVID pneumonia X-ray images independently for learning their respective distributions. As for preparing the data for training the CIN module, for a given image ***X***, we use our trained AEs in the TFEN module and construct the respective output tensor *χ*° according to (5)-(7). We then train our classification model using the output tensor of residual images provided by the TFEN module (as illustrated in Fig. 1).

During the inference, for a new test Chest X-ray image, we first feed this image into the TFEN module and obtain the residual tensor *χ*°, and then, we feed-forward *χ*° into the CIN module to identify the underlying class of the input image. From an interpretability point of view, the overall flow of information can be seen as follows: Given an X-ray image, we consider the closeness of this image to the Healthy, and non-COVID pneumonia as a basis distribution, then, we construct a tensor of residual images that provides a notion of distance of the image and the underlying classes of Healthy and non-COVID. Then, the constructed measures are used for classification purposes.

### 3.3 Quantitative Results

To validate the effectiveness of our proposed model, we quantified the predicted scores of the proposed method using a range of different evaluation metrics including average precision, average recall, average F1-score, sensitivity and the positive predictive value (PPV) for the whole dataset as well as individual categories. Note that for imbalanced classification problems (like ours) where the number of Healthy and non-COVID Pneumonia cases are far more than COVID-19 cases, accuracy is not a good metric and it is essential to evaluate the model performance using precision and recall.

The proposed methodology obtained an overall class average accuracy of 93.5% (Table 2). The high average precision, recall and F1-score, shows that our model performance resulted in lower false positives and false negatives across the whole dataset. The confusion matrix further validates our model’s performance where we achieve a higher *True Positives* for all individual categories (Table 5). In particular, just one COVID-19 case was misclassified into another category. These results are practically very important as misclassifying non-COVID Pneumonia or COVID-19 patients to Healthy category and releasing them can put the mass population at high risk. The sensitivity (Table 3) and PPV scores (Table 4) for individual categories further validates the performance of our model. Moreover, we achieve, comparatively, higher sensitivity and PPV scores for Healthy and non-COVID Pneumonia categories. These results can be attributed to the efficient learning of their individual distributions using the two autoencoders.

**Table 2:** Performance evaluation of the proposed model (%)

| Accuracy | Precision | Recall | F1-score |
| --- | --- | --- | --- |
| 93.50 | 93.63 | 93.50 | 93.51 |

**Table 3:**
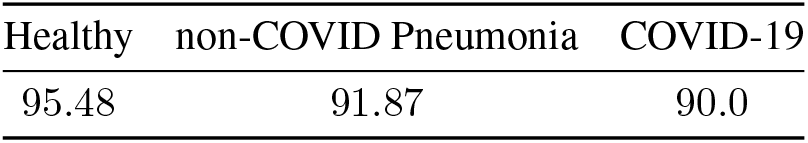
Sensitivity for individual categories (%)

| Healthy | non-COVID Pneumonia | COVID-19 |
| --- | --- | --- |
| 95.48 | 91.87 | 90.0 |

**Table 4:**
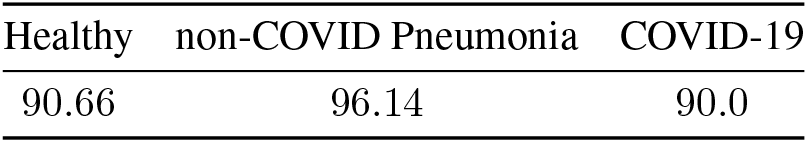
Positive predictive value (PPV) for individual categories (%)

| Healthy | non-COVID Pneumonia | COVID-19 |
| --- | --- | --- |
| 90.66 | 96.14 | 90.0 |

**Table 5:** Confusion matrix for our classification model where we obtain high scores for True Negatives and True Positives. The False Positives for Healthy and non-COVID Pneumonia cases are lower and just one image was misclassified as COVID-19.

|  | Healthy | Pneumonia | COVID-19 |
| --- | --- | --- | --- |
| Healthy | 845 | 39 | 1 |
| Pneumonia | 86 | 972 | 0 |
| COVID-19 | 1 | 0 | 9 |

Last but not the least, the proposed semi-supervised approach results in light-weight deep architecture for screening the COVID-19 from chest X-ray images, with a combined total number of 11.8M trainable parameters which is ∼10 times less parameters than its counterpart proposed in [12], referred to as COVID-Net. Hence, the proposed deep architecture has more potential to be deployed on portable devices with limited processing capabilities.

### 3.4 Prediction Explanation

With the recent emergence of machine learning solutions, in particular deep learning models, it is becoming increasingly important to explain a classifier model’s decision given a certain input. A plethora of explanation algorithms have been proposed for visually explaining an image classifier’s decisions using an attribution map, i.e., a heatmap that highlights the input pixels that are the evidence for or against the classification outputs. Agarwal et al. [15] showed harnessing a generative model for removing features in input images leads to better attribution maps. Due to lack of good generative models for Chest X-Ray images, we chose one of the widely used perturbation-based algorithms for generating attribution maps. Meaningful perturbations are proposed in [22] to learn a minimal salient continuous mask that blurs out an input image in a way that would minimize the target-class probability. For our given classification model, we followed the work by Fong et al. [22] and optimized a small mask of size 28 × 28 and upsampled it to the image size. In Figure 2, we show the attribution maps of some randomly chosen Chest X-ray images from each category of our testing dataset.

**Figure 2:**
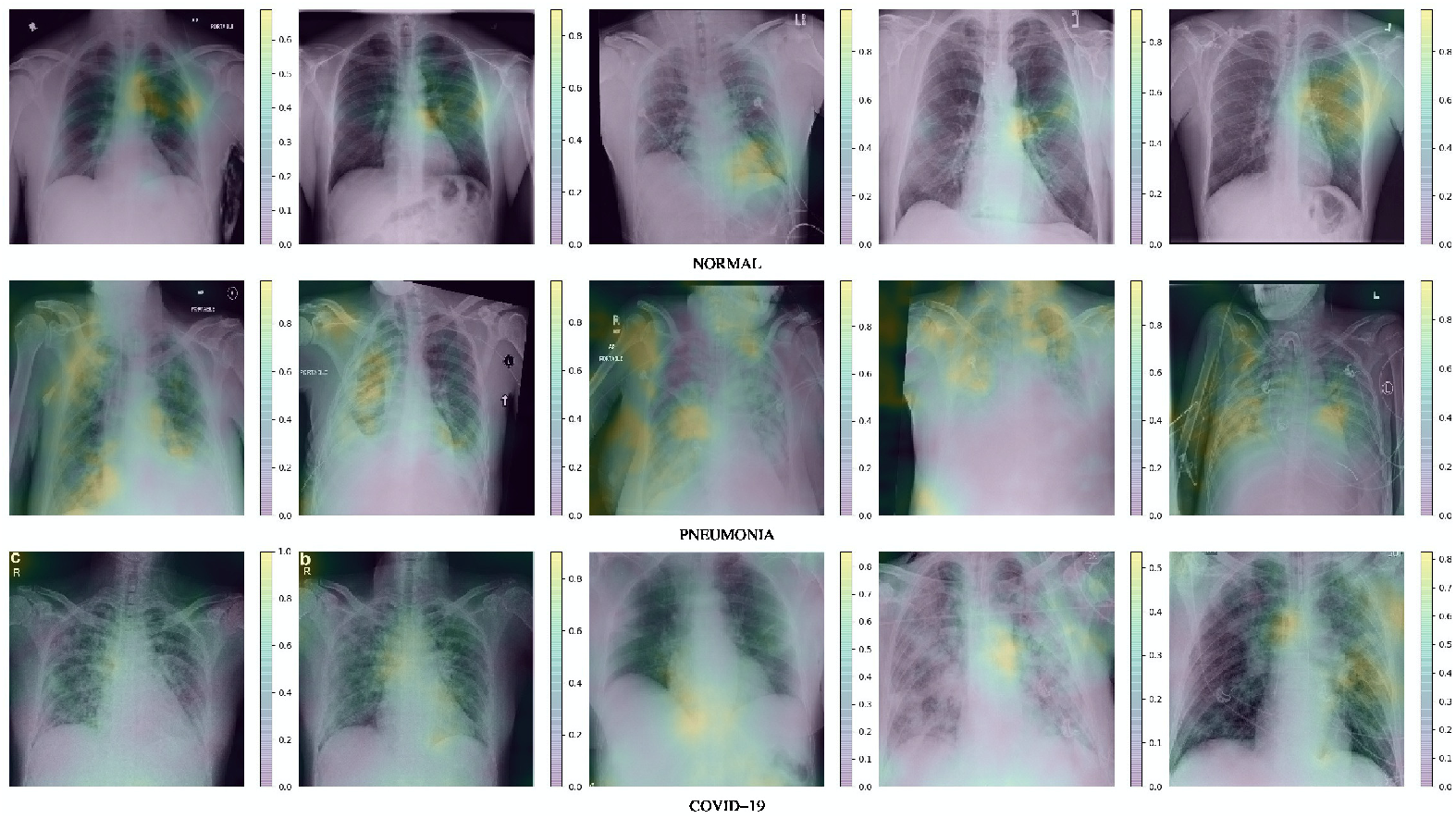
Attribution maps for 5 random patients using our classification model for Healthy, non-COVID Pneumonia, and COVID-19 patients. The colobar map in the respective image represent the salient regions, i.e., yellow means areas that are most salient and blue represents less salient. The explanation maps highlight that our model was consistent in classifying Healthy, non-COVID Pneumonia, and COVID-19 patients by ‘looking’ at different region of the Chest X-rays. Interestingly, most Pneumonia and COVID-19 cases highlights a more diffuse ‘interstitial’ pattern in both lungs.

The attribution maps show that our classification model focuses on different regions in the Chest X-ray images for categorizing the images to Normal, non-COVID Pneumonia, and COVID-19 patients.

## 4 Discussion and Concluding Remarks

In this paper, we considered the identification of COVID-19 cases from X-ray images and proposed a novel semi-supervised deep architecture that can distinguish between the three cases of Healthy, non-COVID pneumonia, COVID-19 infection based on the chest X-ray manifestation of these classes. The proposed methodology is comprised of two modules: 1) the Task-Based Feature Extraction Network (TFEN), and 2) the COVID-19 Identification Network (CIN). The TFEN module is trained based on a semi-supervised methodology and makes use of two autoencoders that allows for automatic segmentation of the infected regions from a latent representation viewpoint. Furthermore, the TFEN module allows for capturing the unique features of each class for later use in the CIN module. Next, the output of the TFEN module is fed into a powerful classifier to provide a classification task on the underlying data. Overall, the proposed methodology is a task-specific deep model and can be seen as an amalgamation of semi-supervised feature extraction methodology, transfer learning, and supervised classification techniques. Our numerical investigations demonstrate that the proposed model achieves superior performance as compared to the state-of-the-art in terms of accuracy, sensitivity, and predicted positive value for the task of classification of COVID-19 from chest X-ray images when the training data-points are scarce. Last but not the least, the proposed network has ∼10 times less trainable parameters compared to existing works (e.g., the COVID-Net [12]), and hence, is very light-weight as compared to the existing bulky networks for screening of COVID-19 using radiography imaging—making it a great candidate for deployment on portable or cheaper devices. To the best of our knowledge, this is the first work that considers a semi-supervised approach to COVID-19 diagnosis with the best identification performance reported to date.

## Data Availability

We have made the codes of our proposed framework publicly available to the research and healthcare community. The codes are available at: https://github.com/chirag126/CoroNet

## Footnotes

1 The codes are available at: https://github.com/chirag126/CoroNet

## References

[1] Ying Song, Shuangjia Zheng, Liang Li, Xiang Zhang, Xiaodong Zhang, Ziwang Huang, Jianwen Chen, Huiying Zhao, Yusheng Jie, Ruixuan Wang, et al. Deep learning enables accurate diagnosis of novel coronavirus (COVID-19) with CT images. medRxiv, 2020.

[2] Weifang Kong and Prachi P Agarwal. Chest imaging appearance of COVID-19 infection.Radiology: Cardiothoracic Imaging, 2(1):e200028, 2020.

[3] Xiaowei Xu, Xiangao Jiang, Chunlian Ma, Peng Du, Xukun Li, Shuangzhi Lv, Liang Yu, Yanfei Chen, Junwei Su, Guanjing Lang, et al. Deep learning system to screen coronavirus disease 2019 pneumonia. arXiv preprint 2002.09334, 2020.

[4] Shahin Khobahi, Naveed Naimipour, Mojtaba Soltanalian, and Yonina C Eldar. Deep signal recovery with one-bit quantization. In IEEE International Conference on Acoustics, Speech and Signal Processing (ICASSP), pages 2987–2991. IEEE, 2019.

[5] Shahin Khobahi and Mojtaba Soltanalian. Model-aware deep architectures for one-bit compressive variational autoencoding. arXiv preprint 1911.12410, 2019.

[6] Chirag Agarwal, Shahin Khobahi, Arindam Bose, Mojtaba Soltanalian, and Dan Schonfeld. Deep-URL: A model-aware approach to blind deconvolution based on deep unfolded Richardson-Lucy network. arXiv preprint 2002.01053, 2020.

[7] Naveed Naimipour, Shahin Khobahi, and Mojtaba Soltanalian. Upr: A model-driven architecture for deep phase retrieval. arXiv preprint 2003.04396, 2020.

[8] Fei Shan+, Yaozong Gao+, Jun Wang, Weiya Shi, Nannan Shi, Miaofei Han, Zhong Xue, Dinggang Shen, and Yuxin Shi. Lung infection quantification of covid-19 in ct images with deep learning. arXiv preprint 2003.04655, 2020.

[9] Shuai Wang, Bo Kang, Jinlu Ma, Xianjun Zeng, Mingming Xiao, Jia Guo, Mengjiao Cai, Jingyi Yang, Yaodong Li, Xiangfei Meng, et al. A deep learning algorithm using ct images to screen for corona virus disease (covid-19). medRxiv, 2020.

[10] Christian Szegedy, Wei Liu, Yangqing Jia, Pierre Sermanet, Scott Reed, Dragomir Anguelov, Dumitru Erhan, Vincent Vanhoucke, and Andrew Rabinovich. Going deeper with convolutions. In Proceedings of the IEEE conference on computer vision and pattern recognition, pages 1–9, 2015.

[11] Ali Narin, Ceren Kaya, and Ziynet Pamuk. Automatic detection of coronavirus disease (covid-19) using x-ray images and deep convolutional neural networks. arXiv preprint 2003.10849, 2020.

[12] Linda Wang and Alexander Wong. Covid-net: A tailored deep convolutional neural network design for detection of covid-19 cases from chest radiography images. arXiv preprint 2003.09871, 2020.

[13] Joseph Paul Cohen, Paul Morrison, and Lan Dao. Covid-19 image data collection. 2003.11597, 2020.

[14] RSNA Pneumonia Detection Challenge. Radiological society of north america, 2018.

[15] Chirag Agarwal, Dan Schonfeld, and Anh Nguyen. Removing input features via a generative model to explain their attributions to classifier’s decisions. arXiv preprint 1910.04256, 2019.

[16] Xiaoran Chen and Ender Konukoglu. Unsupervised detection of lesions in brain mri using constrained adversarial auto-encoders. arXiv preprint 1806.04972, 2018.

[17] Thomas Schlegl, Philipp Seeböck, Sebastian M Waldstein, Ursula Schmidt-Erfurth, and Georg Langs. Unsupervised anomaly detection with generative adversarial networks to guide marker discovery. In International conference on information processing in medical imaging, pages 146–157. Springer, 2017.

[18] Jia Deng, Wei Dong, Richard Socher, Li-Jia Li, Kai Li, and Li Fei-Fei. Imagenet: A large-scale hierarchical image database. In 2009 IEEE conference on computer vision and pattern recognition, pages 248–255. Ieee, 2009.

[19] Tsung-Yi Lin, Piotr Dollár, Ross Girshick, Kaiming He, Bharath Hariharan, and Serge Belongie. Feature pyramid networks for object detection. In Proceedings of the IEEE conference on computer vision and pattern recognition, pages 2117–2125, 2017.

[20] Diederik P Kingma and Jimmy Ba. Adam: A method for stochastic optimization. arXiv preprint 1412.6980, 2014.

[21] Adam Paszke, Sam Gross, Francisco Massa, Adam Lerer, James Bradbury, Gregory Chanan, Trevor Killeen, Zeming Lin, Natalia Gimelshein, Luca Antiga, et al. Pytorch: An imperative style, high-performance deep learning library. In Advances in Neural Information Processing Systems, pages 8024–8035, 2019.

[22] Ruth C Fong and Andrea Vedaldi. Interpretable explanations of black boxes by meaningful perturbation. In Proceedings of the IEEE International Conference on Computer Vision, pages 3429–3437, 2017.

